# Impact of dementia, living in a long-term care facility, and physical activity status on COVID-19 severity in older adults

**DOI:** 10.1101/2022.07.01.22277144

**Authors:** Shinya Tsuzuki, Takayuki Akiyama, Nobuaki Matsunaga, Norio Ohmagari

## Abstract

**Background:** Japan is fast becoming an extremely aged society and older adults are known to be at risk of severe COVID-19. However, the impact of risk factors specific to this population for severe COVID-19 caused by the Omicron variant of concern (VOC) are not yet clear.

**Methods:** We performed an exploratory analysis using logistic regression to identify risk factors for severe COVID-19 illness among 4,868 older adults with a positive SARS-CoV-2 test result who were admitted to a healthcare facility between 1 January 2022 and 16 May 2022. We then conducted one-to-one propensity score (PS) matching for three factors—dementia, admission from a long-term care facility, and poor physical activity status—and used Fisher’s exact test to compare the proportion of severe COVID-19 cases in the matched data. We also estimated the average treatment effect on treated (ATT) in each PS matching analysis.

**Results:** Of the 4,868 cases analyzed, 1,380 were severe. Logistic regression analysis showed that age, male sex, cardiovascular disease, cerebrovascular disease, chronic lung disease, renal failure and/or dialysis, physician-diagnosed obesity, admission from a long-term care facility, and poor physical activity status were risk factors for severe disease. Vaccination and dementia were identified as factors associated with non-severe illness. The ATT for dementia, admission from a long-term care facility, and poor physical activity status was −0.04 (95% confidence interval −0.07, −0.01), 0.09 (0.06, 0.12), and 0.17 (0.14, 0.19), respectively.

**Conclusions:** Our results suggest that poor physical activity status and living in a long-term care facility have a substantial impact on the risk of severe COVID-19 caused by the Omicron VOC, while dementia might be associated with non-severe illness.

## Introduction

Coronavirus disease 2019 (COVID-19), caused by severe acute respiratory coronavirus 2 (SARS-CoV-2), has become a global health threat [1–3]. Since the first case of COVID-19 was identified in Wuhan, China [4], there have been various changes in the approaches to clinical management of COVID-19. The risk factors for severe disease now seem to have been identified [5,6] and the pharmaceutical treatments for COVID-19 cases are at least somewhat established [7–9]. However, the SARS-CoV-2 B.1.1.529 variant of concern (VOC) that causes COVID-19 has somewhat different clinical characteristics from the pre-existing variants. This variant, which was first identified in South Africa on 24 November 2021 [10] and was subsequently named Omicron, has now spread worldwide. Fewer patients develop serious illness with this variant and vaccines are less effective [11–13].

In Japan, the Omicron VOC has spread rapidly, as in many countries, although there have been fewer severe cases than when the Delta variant was dominant [14]. Nevertheless, the higher infectivity of the Omicron VOC than for previous variants led to a huge number of new confirmed cases and a corresponding increase in the number of severe cases. This has burdened our health systems and society further [15].

Although previous studies have identified the risk factors associated with severe illness caused by other variants [5,16–18], the same factors may not be applicable to the disease caused by Omicron VOC. Examining the risk factors associated with severe COVID-19 caused by the Omicron VOC is therefore desirable especially since, throughout the entire pandemic period, older Japanese adults have accounted for a large proportion of the severe COVID-19 cases that required hospitalization [19,20]. Because Japan is fast becoming an extremely aged society, an evaluation of the risk factors specific to the elderly population would be extremely valuable. For instance, the US Centers for Disease Control and Prevention (CDC) reported that living in a long-term care facility was an independent risk factor for mortality [21]. Moreover, dementia or pre-existing Alzheimer’s disease was reported to be associated with late mortality due to COVID-19 [22,23]. According to Steenkamp et al., a moderate or high level of physical activity had a preventive effect on severe COVID-19 [24]. However, the impact of these factors on the severity of COVID-19 caused by the Omicron VOC is not clear.

The main objectives of this study were to identify the risk factors for severe COVID-19 caused by the Omicron VOC and to assess the impact of three factors specific to the elderly population—dementia, living in a long-term care facility, and physical activity status—on the severity of the disease.

## Methods

### Study population and data

Healthcare facilities that voluntarily participated in the COVID-19 Registry Japan (COVIREGI-JP) [19,25], which is managed by the REBIND (Repository of Data and Biospecimen of Infectious Disease) project, enrolled the patients. Research collaborators in each facility manually entered the data into the registry. The inclusion criteria for enrolment were (i) a positive SARS-CoV-2 test result and (ii) admittance to a healthcare facility between 1 January 2022 and 16 May 2022. The exclusion criteria were positive test results for any of the N501Y, E484K, E484Q, and L452R mutants in SARS-CoV-2 genome tests.

### Statistical analysis

As a descriptive analysis of the whole data, continuous variables are presented as median and interquartile range (IQR) and categorical variables as number of cases and percentages. We then performed an exploratory analysis using logistic regression to identify risk factors for severe illness. In this study, we defined severe illness as a need for supplementary oxygen during admission. The following variables were included in the regression model: age, sex, vaccinated at least twice, current smoking habit, cardiovascular disease, cerebrovascular disease, chronic lung disease, asthma, liver disease, renal failure and/or dialysis, diabetes mellitus, solid tumor, blood cancer, collagen disease, physician-diagnosed obesity, dementia, admission from a long-term care facility, and physical activity. We defined physical activity as a dichotomous variable and patients were considered to have good physical activity status if they could (i) eat a normal diet, (ii) walk independently, and (iii) take care of themselves; otherwise, they were considered to have poor physical activity status. Each of the three criteria was assessed by a physician.

Next, we conducted one-to-one propensity score (PS) matching [26] using three categorizations: presence/absence of dementia, admission from a long-term care facility/home or an acute healthcare facility; and good/poor physical activity status. The PS was calculated by logistic regression analysis with the same variables included in the exploratory analysis. All matching processes were based on nearest neighbor matching with a caliper width of 0.05 and no replacement was allowed. The standardized difference was used to measure the covariate balance, and an absolute standardized difference above 0.1 was interpreted as a meaningful imbalance. Fisher’s exact test was used to compare the proportion of severe COVID-19 cases in matched data. In addition, we estimated the average treatment effect on treated (ATT) in each PS matching analysis.

Two-sided *p* values of < 0.05 were considered to show statistical significance. All analyses were conducted with R version 4.1.3 [27].

## Results

The characteristics of participants are shown in Table 1. We included 4,868 patients in the analysis, 1,380 of whom had severe COVID-19. The age distribution of the severe group was about 20 years older than that of the non-severe group. The severe group showed a higher proportion of past medical history and comorbidity.

Table 2 and Figure 1 show the results of the logistic regression analysis. Age, male sex, cardiovascular disease, cerebrovascular disease, chronic lung disease, renal failure and/or dialysis, physician-diagnosed obesity, admission from a long-term care facility, and poor physical activity status were identified as risk factors for severe illness (i.e., need for supplementary oxygen during admission). Vaccination and dementia were identified as factors associated with non-severe illness.

**Figure 1.**
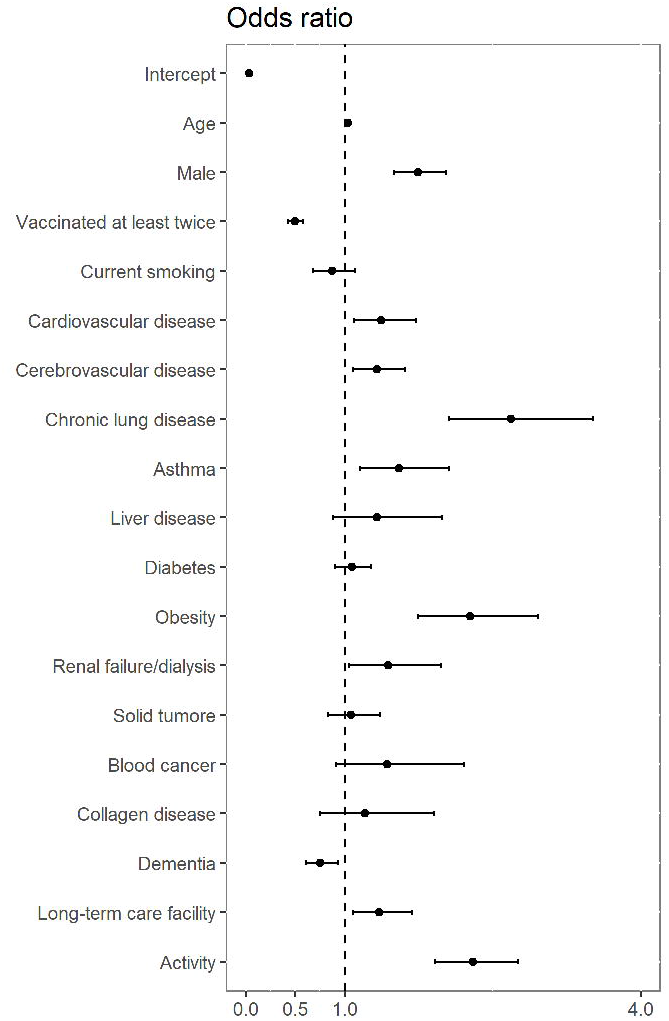
Results of multivariable logistic regression analysis. Black circles indicate median. Whiskers indicate 95% confidence intervals. LTCF, long-term care facility.

Figures 2, 3, and 4 show the standardized mean difference before and after the matching procedure for dementia, admission from a long-term care facility, and physical activity status, respectively. For all categorizations, neither group showed significant differences in each item included in the model. The details of the three datasets after matching are available in Tables S1, S2, and S3 in the supplementary file.

**Figure 2.**
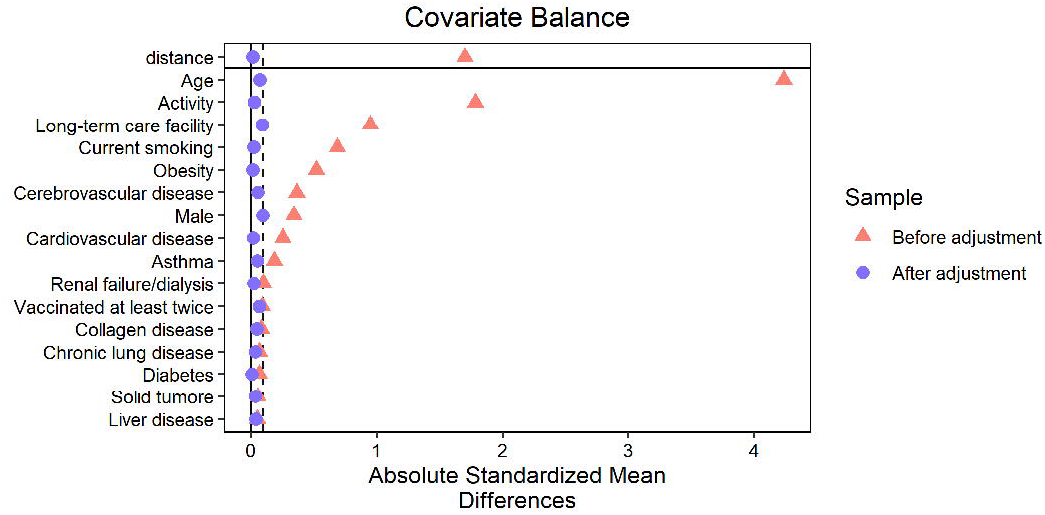
Balance of demographic characteristics of older COVID-19 inpatients before and after propensity score matching (with dementia/without dementia) LTCF, long-term care facility.

**Figure 3.**
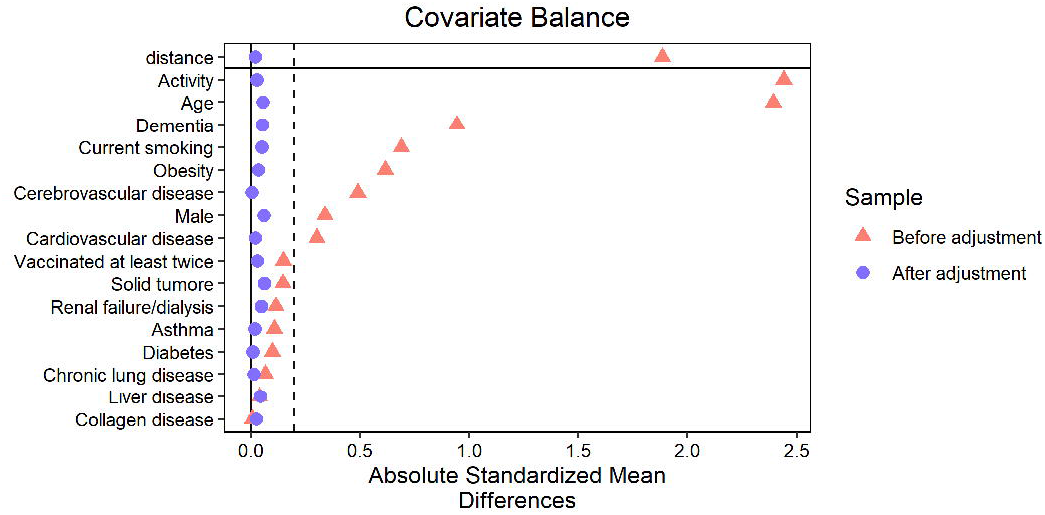
Balance of patients’ demographic characteristics before and after propensity score matching (admission from a long-term care facility/elsewhere)

**Figure 4.**
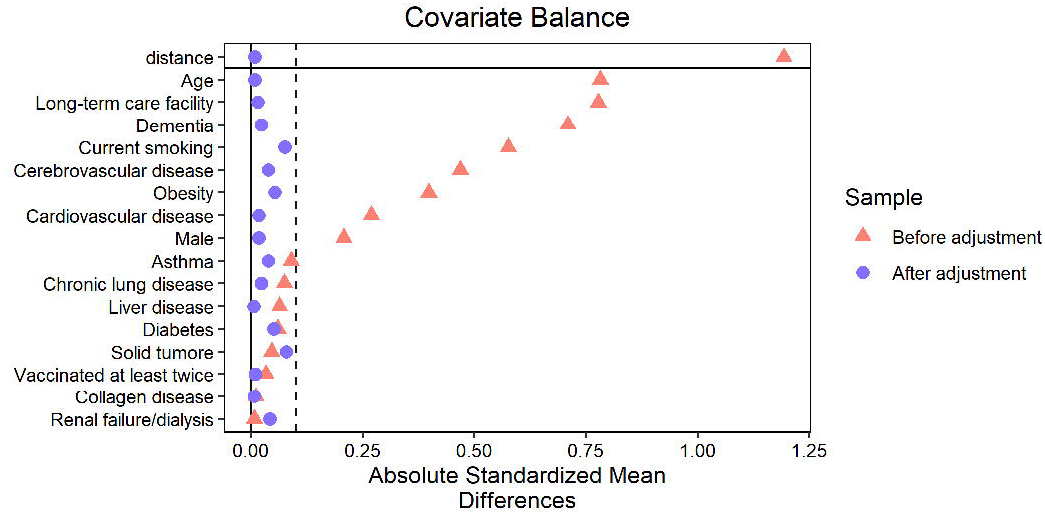
Balance of patients’ demographic characteristics before and after propensity score matching (poor/good physical activity status) LTCF, long-term care facility.

Table 3 shows the estimated ATT of each cohort. All three factors were significantly associated with disease severity. Dementia showed a negative impact on illness severity. Both admission from a long-term care facility and poor physical activity status were associated with severe illness, with the latter showing a larger ATT. Fisher’s exact test determined similar results for admission from a long-term care facility and poor physical activity status (*p* = 0.014 and < 0.001, respectively), whereas dementia did not show a significant difference (*p* = 0.435).

## Discussion

This study identified the risk factors for severe illness due to COVID-19 caused by the Omicron VOC in older adults. These is little evidence on these factors from Japan until now, and our results show that the risk factors for severe illnesses due to the Omicron VOC are very similar to those of other variants (e.g., Alpha and Delta variants). As suggested previously, older age, male sex, cardiovascular disease, chronic lung disease, and obesity are among the factors associated with severe COVID-19 [5,9,12,17,18]. In addition, these risk factors are similar to those specific to the elderly population [6]. These facts may suggest that the original pathology of SARS-CoV-2 infection is inflammation of the respiratory tract, regardless of the strain or variant of the causative organism.

At least two vaccine doses is strongly associated with non-severe illness for the Omicron VOC, as well as for other strains [13,28–30]. This result supports the need to promote vaccination at the population level, even though there has been concern about the efficacy of the currently available COVID-19 vaccines against the Omicron VOC [13].

The results pertaining to dementia should be carefully interpreted. Although Fisher’s exact test did not show significant differences in the risk of severe disease in the data after PS matching, logistic regression analysis identified dementia as one of the risk factors for severe illness. In addition, the ATT for dementia showed a negative impact on its severity. These results are not in agreement with those of previous studies [22,23,31,32], and dementia itself would not intuitively seem to have a positive effect on the severity of COVID-19. However, two reports from Japan stated that dementia was not a significant risk factor for severe illness due to COVID-19 [6,33]. Examination of the reason for this discrepancy is a future challenge.

According to our results, both living in a long-term care facility and poor physical activity status were associated with severe COVID-19, but the ATT for poor physical activity status was the larger of the two. This may suggest that poor physical activity status has a greater impact on the severity of COVID-19 than living in a long-term care facility; in other words, older adults who live in such facilities may also have different risk. Those who have a good physical activity status may have a lower risk of severe illness even if they live in a long-term care facility. This finding may be beneficial when we consider how long-term care facilities manage COVID-19, particularly in super-aged societies like Japan.

Several limitations of this study should be noted. First, this was a retrospective cohort study and not a randomized controlled trial. Although we adjusted for various factors using PS matching, not all factors were included in the model. Next, we excluded a large number of patients from the final analyses because if any two groups (i.e., presence/absence of dementia, living in a long-term care facility/elsewhere, good/poor physical activity status) had substantially different demographic characteristics, then numerous patients had to be removed to adjust the background of both groups. Although this would strengthen the internal validity of our results, the external validity and generalizability were sacrificed to some extent. Furthermore, because our registry data did not include information directly assessing the activities of daily living of each patient (e.g., Barthel Index), we had to adopt a new indicator comprising several factors associated with physical activity. These factors—normal diet, independent walking, and ability to take care of oneself—were assessed by each physician in each facility participating in our registry, and subjectivity might have affected the result.

## Conclusions

Our results suggest that physical activity and living in a long-term care facility have a substantial impact on severe illness caused by COVID-19 owing to the Omicron-19 variant, whereas dementia might be associated with non-severe illness. We should take these factors into consideration in the management of older adults with COVID-19.

## Data Availability

The data supporting the findings of this study are not publicly available due to the privacy of research participants and sites but are available upon reasonable request. Data on an individual level are shared with limitations to participating healthcare facilities through application to the REBIND project.

## Acknowledgments

The authors thank all of the participating facilities for their care of patients with COVID-19 and their cooperation with data entry into the registry. The data used for this research were provided by the REBIND (Repository of Data and Biospecimen of Infectious Disease) project, which was commissioned for the National Center for Global Health and Medicine by the Ministry of Health, Labour and Welfare of Japan.

## Ethics approval

Our study data were provided by Research Electronic Data Capture, a secure, Web-based data capture application hosted at the JCRAC Data Center of the National Center for Global Health and Medicine. The opt-out recruitment method was applied, and informed consents for individuals were waived, as approved by the National Center for Global Health and Medicine Ethics Review Board. Information about the entire research is available through the COVID-19 Registry Japan website (https://covid-registry.ncgm.go.jp/). This study was approved by the National Center for Global Health and Medicine Ethics Review Board (approval number: NCGM-G-003494-0).

## Authors’ contributions

ST and NO conceived the study. TA curated the data. ST and TA analyzed and interpreted the data. ST wrote the first draft of the manuscript. All authors critically reviewed the manuscript and approved the final version.

