## Supplementary material for "Impact of dementia, living in a long-term care facility, and physical activity status on COVID-19 severity in older adults": Tables

**Table 1. Demographic characteristics of older adults hospitalized for COVID-19 caused by the Omicron variant of concern**

| **Supplementary oxygen during admission** | **No** | **Yes** | ***P* value** |
| --- | --- | --- | --- |
| Number of cases | 3488 | 1380 |  |
| Age [median, IQR] | 57.0 [31.0, 78.0] | 79.0 [68.0, 87.0] | <0.001 |
| Male sex (%) | 1693 (48.5) | 802 (58.1) | <0.001 |
| Vaccinated at least twice (%) | 2267 (65.0) | 843 (61.1) | 0.011 |
| Booster dose (%) | 314 (9.0) | 121 (8.8) | 0.824 |
| Current smoking (%) | 442 (12.7) | 117 (8.5) | <0.001 |
| Cardiovascular disease (%) | 202 (5.8) | 198 (14.3) | <0.001 |
| Cerebrovascular disease (%) | 297 (8.5) | 286 (20.7) | <0.001 |
| Chronic lung disease (%) | 120 (3.4) | 172 (12.5) | <0.001 |
| Asthma (%) | 211 (6.0) | 100 (7.2) | 0.135 |
| Liver disease (%) | 73 (2.1) | 51 (3.7) | 0.002 |
| Diabetes mellitus (%) | 591 (16.9) | 344 (24.9) | <0.001 |
| Obesity (%) | 229 (6.6) | 111 (8.0) | 0.071 |
| Renal failure/dialysis (%) | 119 (3.4) | 80 (5.8) | <0.001 |
| Solid tumor (%) | 227 (6.5) | 138 (10.0) | <0.001 |
| Blood cancer (%) | 70 (2.0) | 38 (2.8) | 0.130 |
| Collagen disease (%) | 71 (2.0) | 35 (2.5) | 0.278 |
| Dementia (%) | 349 (10.0) | 315 (22.8) | <0.001 |
| Living in LTCF (%) | 311 (8.9) | 357 (25.9) | <0.001 |
| Normal diet (%) | 3045 (87.3) | 929 (67.3) | <0.001 |
| Independent walking (%) | 2813 (80.6) | 723 (52.4) | <0.001 |
| Self-care ability (%) | 2802 (80.3) | 710 (51.4) | <0.001 |
| Poor physical activity status (%) | 800 (22.9) | 728 (52.8) | <0.001 |
| Outcome |  |  | NA |
| Discharged to home (%) | 2732 (78.3) | 664 (48.1) |  |
| Isolated (%) | 267 (7.7) | 16 (1.2) |  |
| Discharged to LTCF (%) | 348 (10.0) | 310 (22.5) |  |
| Transfer (%) | 130 (3.7) | 183 (13.3) |  |
| Transfer to higher-level facility (%) | 2 (0.1) | 21 (1.5) |  |
| Death (%) | 9 (0.3) | 181 (13.1) |  |
| Others (%) | 0 (0.0) | 5 (0.4) |  |

IQR, interquartile range; LTCF, long-term care facility; NA, not applicable

**Table 2. Results of logistic regression analysis**

| **Variable** | **Odds ratio** | **95% confidence interval** | ***P* value** |
| --- | --- | --- | --- |
| Intercept | −3.40 | [−3.69, −3.13] | <0.001 |
| Age | 0.03 | [0.03, 0.04] | <0.001 |
| Male sex | 0.55 | [0.40, 0.71] | <0.001 |
| Vaccinated at least twice | −0.70 | [−0.85, −0.55] | <0.001 |
| Current smoking | −0.13 | [−0.38, 0.10] | 0.272 |
| Cardiovascular disease | 0.32 | [0.09, 0.55] | 0.007 |
| Cerebrovascular disease | 0.28 | [0.08, 0.48] | 0.006 |
| Chronic lung disease | 0.99 | [0.72, 1.26] | <0.001 |
| Asthma | 0.44 | [0.15, 0.72] | 0.003 |
| Liver disease | 0.28 | [−0.13, 0.68] | 0.170 |
| Diabetes mellitus | 0.07 | [−0.10, 0.24] | 0.411 |
| Obesity | 0.82 | [0.55, 1.09] | <0.001 |
| Renal failure/dialysis | 0.37 | [0.04, 0.68] | <0.025 |
| Solid tumor | 0.06 | [−0.19, 0.31] | 0.622 |
| Blood cancer | 0.36 | [−0.09, 0.79] | 0.110 |
| Collagen disease | 0.18 | [−0.29, 0.64] | 0.435 |
| Dementia | −0.28 | [−0.50, −0.07] | 0.011 |
| Living in LTCF | 0.30 | [0.08, 0.52] | 0.007 |
| Poor physical activity status | 0.83 | [0.65, 1.01] | <0.001 |

LTCF, long-term care facility

**Table 3. Average treatment effect on the treated for each matched cohort**

| Factor | ATT | 95% confidence interval | *P* value |
| --- | --- | --- | --- |
| Dementia | −0.04 | [−0.07, −0.01] | 0.004 |
| Living in LTCF | 0.09 | [0.06, 0.12] | <0.001 |
| Poor physical activity status | 0.17 | [0.14, 0.19] | <0.001 |

ATT, average treatment effect on the treated; LTCF, long-term care facility
