## Supplementary file for "Impact of dementia, living in a long-term care facility, and physical activity status on COVID-19 severity in older adults"

**Supplementary Table S1. Demographic characteristics of hospitalized older adults with COVID-19 caused by the Omicron variant of concern after propensity score matching (with dementia/without dementia)**

| **Dementia** | **No** | **Yes** | ***P* value** | **SMD** |
| --- | --- | --- | --- | --- |
| Number of cases | 555 | 555 |  |  |
| Age [median, IQR] | 86.0 [82.0, 91.0] | 86.0 [81.0, 90.0] | 0.360 | 0.070 |
| Male sex (%) | 245 (44.1) | 220 (39.6) | 0.144 | 0.091 |
| Vaccinated at least twice (%) | 396 (71.4) | 379 (68.3) | 0.295 | 0.067 |
| Current smoking (%) | 12 (2.2) | 14 (2.5) | 0.843 | 0.024 |
| Cardiovascular disease (%) | 95 (17.1) | 91 (16.4) | 0.810 | 0.019 |
| Cerebrovascular disease (%) | 140 (25.2) | 153 (27.6) | 0.414 | 0.053 |
| Chronic lung disease (%) | 48 (8.6) | 43 (7.7) | 0.662 | 0.033 |
| Asthma (%) | 25 (4.5) | 20 (3.6) | 0.543 | 0.046 |
| Liver disease (%) | 9 (1.6) | 12 (2.2) | 0.661 | 0.040 |
| Diabetes mellitus (%) | 127 (22.9) | 125 (22.5) | 0.943 | 0.009 |
| Obesity (%) | 8 (1.4) | 9 (1.6) | 1.000 | 0.015 |
| Renal failure/dialysis (%) | 15 (2.7) | 17 (3.1) | 0.858 | 0.022 |
| Solid tumor (%) | 44 (7.9) | 39 (7.0) | 0.648 | 0.034 |
| Blood cancer (%) | 14 (2.5) | 7 (1.3) | 0.185 | 0.093 |
| Collagen disease (%) | 12 (2.2) | 9 (1.6) | 0.661 | 0.040 |
| Long-term care facility (%) | 231 (41.6) | 256 (46.1) | 0.147 | 0.091 |
| Activity (%) | 464 (83.6) | 459 (82.7) | 0.748 | 0.024 |
| Supplementary oxygen during admission | 276 (49.7) | 251 (45.2) | 0.149 | 0.090 |

IQR, interquartile range; SMD, standardized mean difference

**Supplementary Table S2. Patients’ demographic characteristics after propensity score matching (admission from a long-term care facility/elsewhere)**

| **Admission from a long-term care facility** | **No** | **Yes** | ***P* value** | **SMD** |
| --- | --- | --- | --- | --- |
| Number of cases | 531 | 531 |  |  |
| Age [median, IQR] | 86.0 [80.0, 90.0] | 86.0 [79.0, 91.0] | 0.952 | 0.055 |
| Male sex (%) | 232 (43.7) | 217 (40.9) | 0.385 | 0.057 |
| Vaccinated at least twice (%) | 368 (69.3) | 361 (68.0) | 0.692 | 0.028 |
| Current smoking (%) | 20 (3.8) | 16 (3.0) | 0.612 | 0.042 |
| Cardiovascular disease (%) | 84 (15.8) | 88 (16.6) | 0.803 | 0.020 |
| Cerebrovascular disease (%) | 159 (29.9) | 160 (30.1) | 1.000 | 0.004 |
| Chronic lung disease (%) | 44 (8.3) | 46 (8.7) | 0.912 | 0.014 |
| Asthma (%) | 25 (4.7) | 27 (5.1) | 0.887 | 0.017 |
| Liver disease (%) | 17 (3.2) | 21 (4.0) | 0.621 | 0.041 |
| Diabetes mellitus (%) | 128 (24.1) | 126 (23.7) | 0.943 | 0.009 |
| Obesity (%) | 10 (1.9) | 8 (1.5) | 0.813 | 0.029 |
| Renal failure/dialysis (%) | 20 (3.8) | 16 (3.0) | 0.612 | 0.042 |
| Solid tumor (%) | 36 (6.8) | 29 (5.5) | 0.443 | 0.055 |
| Blood cancer (%) | 10 (1.9) | 4 (0.8) | 0.177 | 0.099 |
| Collagen disease (%) | 13 (2.4) | 15 (2.8) | 0.849 | 0.024 |
| Dementia (%) | 232 (43.7) | 246 (46.3) | 0.423 | 0.053 |
| Poor activity (%) | 476 (89.6) | 472 (88.9) | 0.766 | 0.024 |
| Supplementary oxygen during admission | 246 (46.3) | 287 (54.0) | 0.014 | 0.155 |

IQR, interquartile range; SMD, standardized mean difference

**Supplementary Table S3. Patients’ demographic characteristics after propensity score matching (good/poor physical activity status )**

| **Physical activity status** | **Good** | **Poor** | ***P* value** | **SMD** |
| --- | --- | --- | --- | --- |
| Number of cases | 819 | 819 |  |  |
| Age [median, IQR] | 73.0 [53.0, 83.0] | 79.0 [52.5, 87.0] | <0.001 | 0.009 |
| Male sex (%) | 440 (53.7) | 433 (52.9) | 0.766 | 0.017 |
| Vaccinated at least twice (%) | 489 (59.7) | 485 (59.2) | 0.880 | 0.010 |
| Current smoking (%) | 39 (4.8) | 51 (6.2) | 0.233 | 0.064 |
| Cardiovascular disease (%) | 100 (12.2) | 95 (11.6) | 0.760 | 0.019 |
| Cerebrovascular disease (%) | 139 (17.0) | 153 (18.7) | 0.401 | 0.045 |
| Chronic lung disease (%) | 64 (7.8) | 59 (7.2) | 0.708 | 0.023 |
| Asthma (%) | 41 (5.0) | 48 (5.9) | 0.513 | 0.038 |
| Liver disease (%) | 25 (3.1) | 26 (3.2) | 1.000 | 0.007 |
| Diabetes mellitus (%) | 186 (22.7) | 169 (20.6) | 0.337 | 0.050 |
| Obesity (%) | 25 (3.1) | 32 (3.9) | 0.419 | 0.047 |
| Renal failure/dialysis (%) | 48 (5.9) | 41 (5.0) | 0.513 | 0.038 |
| Solid tumor (%) | 107 (13.1) | 89 (10.9) | 0.196 | 0.068 |
| Blood cancer (%) | 27 (3.3) | 16 (2.0) | 0.121 | 0.084 |
| Collagen disease (%) | 20 (2.4) | 19 (2.3) | 1.000 | 0.008 |
| Dementia (%) | 96 (11.7) | 105 (12.8) | 0.547 | 0.033 |
| Long-term care facility (%) | 57 (7.0) | 63 (7.7) | 0.636 | 0.028 |
| Supplementary oxygen during admission | 213 (26.0) | 349 (42.6) | <0.001 | 0.355 |

IQR, interquartile range; SMD, standardized mean difference
